# Causal association of the brain structure with the risk of knee osteoarthritis: A large-scale genetic correlation study

**DOI:** 10.1101/2023.12.20.23300318

**Authors:** Zhe Ruan, Shaohai Lin, Zhi Liu, Peng Chen, Tongtong Xie, Li Meng, Haitao Long, Shushan Zhao

**Author notes:** Corresponding authors: Shushan Zhao. Haitao Long. These authors contributed equally to this work (Zhe Ruan, Shaohai Lin).

## Abstract

**Objectives:** Observational studies have shown the association between knee osteoarthritis (KOA) and neurological disorders with alterations in brain imaging-derived phenotypes (BIDPs). This study aimed at investigating whether alterations in brain structure are correlated with the occurrence of KOA.

**Methods:** Based on the summary data from two large scale genome-wide association studies (GWASs), we performed a bidirectional two-sample Mendelian randomization (MR) analysis using single-nucleotide polymorphisms (SNPs) as instrumental variables (IVs) to determine the potential causal relationships between KOA and BIDPs.

**Results:** We identified the genetic correlations of 152 BIDPs with KOA using linkage disequilibrium score regression. MR analysis revealed that increased volume but decreased intensity-contrast of bilateral nucleus accumbens (NAc), as well as increased left paracentral area was positively causally associated with KOA risk. For the IDPs of structural connectivity, we identified causal associations between multiple increased DTI parameter indicators of corticospinal tract (CST) and KOA. Inversely, KOA was positively correlated with the thickness and intensity-contrast of the rostral anterior cingulate, as well as the intensity-contrast of caudal anterior cingulate, insula cortex, and the grey matter volume of pallidum.

**Conclusion:** Our study supported bidirectional causal associations between KOA and BIDPs, which may provide new insights into the interaction of KOA with structural alterations in the nervous system.

## 1. Introduction

Osteoarthritis (OA) is the most common musculoskeletal disorder around the world, it is characterized by articular cartilage degeneration, osteophyte formation, and asymmetric joint spaces narrowing, leading to pain and disability[1]. It has been recently estimated that there is an overall prevalence of about 300 million for OA, of which 263 million had knee osteoarthritis (KOA)[2]. With prolonged life expectancy and population aging, the prevalence of KOA is predicted to dramatically increase in the future. Epidemiologic studies have shown that the most common risk factors for KOA include previous joint injuries, older age, obesity, female gender, and anatomical abnormalities of the joints[3].

It has been found that many neurological disorders such as major depressive disorder (MDD)[4, 5], Alzheimer’s disease (AD)[6, 7], Parkinson’s disease (PD)[8], and cerebrovascular dysfunction diseases[9] are closely related to OA and may be comorbidities or risk factors for it. Neuroimaging techniques have been utilized in recent years for the non-invasive detection of central and peripheral nervous system degeneration so as to monitor disease progression and prognosis, and it has also been applied to investigate brain structure, function and metabolism[10, 11]. It is also worth noting that these disorders usually have neurological degeneration to certain extent. Most magnetic resonance-based structural imaging studies suggested that the degree and rate of atrophy in medial temporal structures, such as the hippocampus, can be recognized as diagnostic and progression markers or AD[12]. Another recent structural network study using diffusion tensor imaging (DTI) revealed that lower fractional anisotropy (FA) values of superior longitudinal fasciculus, prefrontal cortex, and parietal lobe in left hemisphere among patients with MDD compared to health controls[13]. However, there are no studies investigating whether brain structure alterations are associated with the occurrence of KOA.

Mendelian randomization (MR) utilizes genetic variants, such as single-nucleotide polymorphisms (SNPs), as instrumental variables (IVs) to determine the potential associations between exposures and disorders, and it circumvents the bias caused by confounding factors in traditional observational studies[14]. With the development of genome-wide association studies (GWAS), a large number of public databases are now available for MR studies to assess potential causal associations between OA and other diseases[15–17]. Here we used SNPs strongly associated with brain imaging-derived phenotypes (BIDPs) and KOA as IVs. We performed a bidirectional two-sample MR analysis to investigate the association between brain structures and KOA risk.

## 2. Materials and Methods

### 2.1 Brain imaging-derived phenotypes (BIDPs) data

A recent GWAS of brain imaging traits was performed on 33,224 individuals of European ancestry (aged between 40 and 80 years) based on data from the UK Biobank **^Error! Reference source not found.^**. Full summary GWAS statistics for the BIDPs are available from the Oxford Brain Imaging Genetics (BIG40) (https://open.win.ox.ac.uk/ukbiobank/big40).

The selected BIDPs are divided into two categories: brain structure (structural MRI) and brain structural connectivity (diffusion MRI, dMRI). The original IDPs for brain structure are T1- or T2-weighted structural images, which are subsequently processed by different algorithms to generate the final traits. The structural brain connectivity IDPs are initially determined by dMRI, which measures the diffusion of water molecules and allows for the indirect mapping of the axon connectivity in the human brain. Details procedures of imaging acquisition and processing of brain MRI scans can be found in previous studies. Overall, we extracted brain structures including 1,451 global and regional anatomical subtypes, including volume, subcortical thickness and superficial area, regional and tissue intensity, cortical gray-white contrast, as well as 675 brain structural connections, including long-range structural connectivity and local microstructure **(Supplementary Table 1)**.

### 2.2 Knee osteoarthritis (KOA) data

Summary statistics for KOA were obtained from Arthritis Research UK Osteoarthritis Genetics (arcOGEN) Consortium cohorts of European descent (http://www.arcogen.org.uk/), which included 11,655 individuals ( 646 cases and 11,009 controls)[18]. Osteoarthritis cases were identified on the basis of clinical evidence of disease to a level requiring joint replacement or radiographic evidence of disease (Kellgren–Lawrence grade ≥2).

### 2.3 Linkage disequilibrium score regression (LDSC) analysis

LD score regression is a reliable and efficient method to estimate the SNP heritability for complex traits and diseases GWAS summary-level results, and has recently been used to determine whether different traits have overlapping genetic backgrounds[19]. LDSC regression was first used to assess the correlation between BIDPs and KOA, which can be inferred from the regression slope. Variants were excluded if the following criteria were met: missing value, missing in Hapmap3, INFO score ≤ 0.9, minor allele frequency (MAF) ≤ 0.01, non-SNPs or rs number duplicates.

### 2.4 Instrumental variable selection

The genetic variants utilized as IVs for MR should be strongly related to the exposure, independent from any confounding factors, and independent from any pathway associated with the outcome, except for the exposure pathway. In our MR analysis, we initially extracted IVs based on the genome-wide significance threshold (*p* < 5 × 10^-8^), followed by calculating the *F* statistics of each SNP to exclude weak IVs with *F* statistics < 10. We then ensured independence among IVs for each exposure by conducting an LD test on each SNP identified as an IV based on individuals with European ancestry from the 1000 Genomes Project with *r^2^* < 0.001 and a window size of 10,000 kb. Finally, in case the target SNPs were not found in the outcome GWAS, we used SNPs with high LD (*r^2^* > 0.8) to substitute.

### 2.5 Mendelian randomization

To analyze the causal association between BIDPs and KOA, we utilized multiple complementary methods, including inverse variance weighted (IVW), MR-Egger, weighted median, simple mode, weighted mode, and Wald ratio methods. The IVW model was used as the major primary statistical method, and the Wald ratio method was used when a genetic variant contained only one genetically related SNP. We assessed the presence of horizontal pleiotropy by using Cochrańs *Q* statistic to test for heterogeneity among IVs, with *p* < 0.05 indicating heterogeneity. If heterogeneity was present, we used the random-effects model for subsequent analyses; otherwise, we used the fixed-effects model. Leave-one-out cross-validation was performed to evaluate the stability of the MR results through sequential exclusion of each IV. In addition, we conducted MR-Egger analysis to identify directional pleiotropy. Funnel plot asymmetry indicated the presence of horizontal pleiotropy. In the current study, we focused on those positive MR analysis results with the number of SNPs greater than or equal to three.

### 2.6 Statistical Analyses

All statistical analyses were performed using the “TwoSampleMR” and “MR-PRESSO” packages of R software (version 3.3). All estimates are reported as two-tailed *p* values with the threshold of significance at 0.05.

## 3. Results

### 3.1 Linkage Disequilibrium Score Regression

We first examined the genetic association of GWAS datasets of 2,126 BIDPs with KOA using LDSC. A total of 152 BIDPs (53 IDPs of brain structure and 99 IDPs of structural connectivity, respectively) showed significant correlations with the risk of KOA **(Supplementary Table 2)**.

### 3.2 Causal effect of BIDPs on KOA

*F*-statistic is used to assess the strength of the association between instrumental variables and exposure factors. All *F*-statistics are greater than 10 (range:29.73-1081.32, with an average value of 51.89), indicating a relatively low likelihood of weak instrument bias for the instrumental variables **(Supplementary Table 3)**.

In the current MR analysis, a total of 60 BIDPs were identified to be causally associated with the risk of KOA (**Table 1**). Among the brain structural IDPs relevant to the nucleus accumbens (NAc), the increased volume in both left (*β* = 0.4893, *p* = 0.0434) and right (*β* = 0.6877, *p* = 0.029) hemisphere was found to be positively causally associated with KOA. However, the increased intensity contrast of NAc in both hemispheres had a negative causal association with KOA (left: *β* = -0.1829, *p* = 0.046; right: *β* = -0.1862, *p* = 0.0322). In addition, the increased paracentral area in the left hemisphere measured by Desikan (*β* = 0.7444, *p* = 0.0248) and DKTatlas (*β* = 0.7398, *p* = 0.0266) algorithms, the left BA4a area (*β* = 0.7274, *p* = 0.029), the total white matter volume (*β* = 0.3356, *p* = 0.0386), the bilateral gray matter volume of putamen (left: *β* = 0.2259, *p* = 0.009; right: *β* = 0.1957, *p* = 0.0293) and cerebellum_crus_I (left: *β* = 0.2991, *p* = 0.012; right: *β* = 0.2984, *p* = 0.0343), and the bilateral cuneus intensity contrast (left: *β* = 0.2257, *p* = 0.046; right: *β* = 0.2807, *p* = 0.0084) were positively correlated with KOA. We also observed significant causal effects of the decreased left circular sulcus volume (*β* = -1.0172, *p* = 0.0173), right superiortemporal area (*β* = - 0.5876, *p* = 0.0363), left inferiortemporal thickness based on Desikan (*β* = -0.695, *p* = 0.0121) and aparc-a2009s (*β* = -0.6405, *p* = 0.024) parcellation, as well as right amygdala intensity contrast (*β* = -0.6031, *p* = 0.0464) on KOA risk (**Table 1**).

**Table 1.** The results or Forward MR (BIDPs we were used as exposures).

| Exposure<br>BIDPs<br>ID | Outcome | IDP short name | KOA vs Control |  |  |  |  |  |
| --- | --- | --- | --- | --- | --- | --- | --- | --- |
| | | | N (SNPs) | $\beta$ | SE | P value | OR | Method |
| 7 | Knee OA (arcOGEN) | IDP_T1_SIENAX_white_normalised_volume | 16 | 0.335593198 | 0.162237867 | 0.038590616 | 1.398769887 | IVW |
| 8 | Knee OA (arcOGEN) | IDP_T1_SIENAX_white_unnormalised_volume | 15 | 0.364103774 | 0.172827997 | 0.035140062 | 1.439223561 | IVW |
| 24 | Knee OA (arcOGEN) | IDP_T1_FIRST_right_accumbens_volume | 4 | 0.687650204 | 0.314952856 | 0.029010539 | 1.989036208 | IVW |
| 126 | Knee OA (arcOGEN) | IDP_T1_FAST_ROIs_L_putamen | 18 | 0.225852679 | 0.086885759 | 0.009338113 | 1.253391001 | IVW |
| 127 | Knee OA (arcOGEN) | IDP_T1_FAST_ROIs_R_putamen | 16 | 0.195681385 | 0.089810251 | 0.029344203 | 1.216139363 | IVW |
| 144 | Knee OA (arcOGEN) | IDP_T1_FAST_ROIs_L_cerebellum_crus_I | 21 | 0.29906601 | 0.118375693 | 0.01152336 | 1.348598641 | IVW |
| 146 | Knee OA (arcOGEN) | IDP_T1_FAST_ROIs_R_cerebellum_crus_I | 13 | 0.298411367 | 0.140955296 | 0.034254426 | 1.34771608 | IVW |
| 181 | Knee OA (arcOGEN) | aseg_global_volume_Optic-Chiasm | 4 | 0.726762749 | 0.296972732 | 0.014395602 | 2.068373912 | IVW |
| 201 | Knee OA (arcOGEN) | aseg_lh_volume_Accumbens-area | 9 | 0.489288172 | 0.242267012 | 0.043422446 | 1.631154705 | IVW |
| 272 | Knee OA (arcOGEN) | HippSubfield_rh_volume_presubiculum-body | 5 | 0.576319605 | 0.236674376 | 0.014888747 | 1.779477185 | IVW |
| 277 | Knee OA (arcOGEN) | HippSubfield_rh_volume_CA3-body | 6 | 0.518745785 | 0.230132289 | 0.024188622 | 1.679919348 | IVW |
| 289 | Knee OA (arcOGEN) | ThalamNuclei_lh_volume_PuI | 3 | 0.517937664 | 0.237806723 | 0.029407721 | 1.678562318 | IVW |
| 448 | Knee OA (arcOGEN) | aparc-DKTatlas_lh_volume_lingual | 6 | 0.63249724 | 0.207642094 | 0.002318357 | 1.882305283 | IVW |
| 547 | Knee OA (arcOGEN) | aparc-a2009s_lh_volume_S-circular-insula-inf | 3 | -1.017220792 | 0.427168845 | 0.017251259 | 0.361598502 | IVW |
| 664 | Knee OA (arcOGEN) | aparc-Desikan_lh_area_paracentral | 3 | 0.744386864 | 0.331556352 | 0.024759836 | 2.105150295 | IVW |
| 711 | Knee OA (arcOGEN) | aparc-Desikan_rh_area_superiortemporal | 3 | -0.631373884 | 0.312334496 | 0.043231443 | 0.531860584 | IVW |
| 786 | Knee OA (arcOGEN) | BA-exvivo_lh_area_BA4a | 4 | 0.727435725 | 0.333183684 | 0.029014655 | 2.069766347 | IVW |
| 824 | Knee OA (arcOGEN) | aparc-DKTatlas_lh_area_paracentral | 3 | 0.739780786 | 0.333685688 | 0.026623171 | 2.095476105 | IVW |
| 967 | Knee OA (arcOGEN) | aparc-a2009s_rh_area_G-oc-temp-med-Lingual | 9 | 0.421148192 | 0.188918775 | 0.025796919 | 1.523710063 | IVW |
| 1028 | Knee OA (arcOGEN) | aparc-Desikan_lh_thickness_inferiortemporal | 4 | -0.695018214 | 0.276983682 | 0.012099164 | 0.499065358 | IVW |
| 1029 | Knee OA (arcOGEN) | aparc-Desikan_lh_thickness_isthmuscingulate | 3 | -0.752642108 | 0.310066749 | 0.015209361 | 0.471120157 | IVW |
| 1084 | Knee OA (arcOGEN) | aparc-Desikan_rh_thickness_supramarginal | 4 | -0.587562051 | 0.280617622 | 0.036276252 | 0.555680355 | IVW |
| 1099 | Knee OA (arcOGEN) | BA-exvivo_lh_thickness_MT | 4 | -0.683857342 | 0.288409946 | 0.017733815 | 0.504666562 | IVW |
| 1214 | Knee OA (arcOGEN) | aparc-a2009s_lh_thickness_G-temporal-inf | 4 | -0.640463217 | 0.283699208 | 0.023974154 | 0.52704823 | IVW |
| 1327 | Knee OA (arcOGEN) | aseg_global_intensity_4th-Ventricle | 5 | -0.554646703 | 0.232468154 | 0.017037197 | 0.574275115 | IVW |
| 1349 | Knee OA (arcOGEN) | aseg_lh_intensity_Accumbens-area | 18 | -0.182872874 | 0.09163022 | 0.04595893 | 0.832874029 | IVW |
| 1362 | Knee OA (arcOGEN) | aseg_rh_intensity_Amygdala | 3 | -0.603116447 | 0.302801552 | 0.046394342 | 0.547103956 | IVW |
| 1363 | Knee OA (arcOGEN) | aseg_rh_intensity_Accumbens-area | 18 | -0.186169809 | 0.086901398 | 0.032168475 | 0.830132619 | IVW |
| 1371 | Knee OA (arcOGEN) | wg_lh_intensity-contrast_cuneus | 18 | 0.225734307 | 0.113240385 | 0.046216772 | 1.253242643 | IVW |
| 1406 | Knee OA (arcOGEN) | wg_rh_intensity-contrast_cuneus | 22 | 0.280719327 | 0.10646927 | 0.008373584 | 1.324081918 | IVW |
| 1439 | Knee OA (arcOGEN) | IDP_SWI_T2star_right_thalamus | 5 | -0.466016192 | 0.222154767 | 0.035930805 | 0.627497124 | IVW |
| 1491 | Knee OA (arcOGEN) | IDP_dMRI_TBSS_FA_Fornix_cres+Stria_terminalis_L | 8 | 0.710499467 | 0.297056239 | 0.016765921 | 2.035007423 | IVW |
| 1603 | Knee OA (arcOGEN) | IDP_dMRI_TBSS_MD_Pontine_crossing_tract | 4 | -1.038511303 | 0.332764364 | 0.001803232 | 0.353981261 | IVW |
| 1651 | Knee OA (arcOGEN) | IDP_dMRI_TBSS_L1_Pontine_crossing_tract | 8 | -0.86702052 | 0.219996619 | 8.11213E-05 | 0.420201668 | IVW |
| 1667 | Knee OA (arcOGEN) | IDP_dMRI_TBSS_L1_Anterior_limb_of_internal_capsule_L | 10 | -0.471261642 | 0.186928873 | 0.011699703 | 0.624214237 | IVW |
| 1696 | Knee OA (arcOGEN) | IDP_dMRI_TBSS_L1_Tapetum_R | 3 | -0.730049887 | 0.322082217 | 0.023411192 | 0.48188495 | IVW |
| 1704 | Knee OA (arcOGEN) | IDP_dMRI_TBSS_L2_Corticospinal_tract_R | 7 | -0.525102523 | 0.25581608 | 0.04010591 | 0.59149472 | IVW |
| 1705 | Knee OA (arcOGEN) | IDP_dMRI_TBSS_L2_Corticospinal_tract_L | 7 | -0.808443389 | 0.264859886 | 0.00227062 | 0.445551076 | IVW |
| 1747 | Knee OA (arcOGEN) | IDP_dMRI_TBSS_L3_Pontine_crossing_tract | 3 | -1.100943965 | 0.385352012 | 0.004276894 | 0.332557013 | IVW |
| 1844 | Knee OA (arcOGEN) | IDP_dMRI_ProbtrackX_L1_str_l | 7 | -0.37913881 | 0.185718772 | 0.041204395 | 0.684450597 | IVW |
| 1845 | Knee OA (arcOGEN) | IDP_dMRI_ProbtrackX_L1_str_r | 6 | -0.484829751 | 0.239481298 | 0.042918791 | 0.615802027 | IVW |
| 1861 | Knee OA (arcOGEN) | IDP_dMRI_ProbtrackX_L2_ifo_r | 13 | -0.291854609 | 0.132295676 | 0.027378524 | 0.746877117 | IVW |
| 1918 | Knee OA (arcOGEN) | IDP_dMRI_TBSS_ICVF_Anterior_limb_of_internal_capsule_R | 15 | 0.283740719 | 0.136324102 | 0.037400171 | 1.328088538 | IVW |
| 2053 | Knee OA (arcOGEN) | IDP_dMRI_TBSS_ISOVF_Pontine_crossing_tract | 4 | -0.897098021 | 0.418801426 | 0.032188608 | 0.40775123 | IVW |
| 2059 | Knee OA (arcOGEN) | IDP_dMRI_TBSS_ISOVF_Corticospinal_tract_L | 4 | -1.077645475 | 0.332915164 | 0.001207946 | 0.340396054 | IVW |
| 2069 | Knee OA (arcOGEN) | IDP_dMRI_TBSS_ISOVF_Anterior_limb_of_internal_capsule_L | 7 | 0.575327735 | 0.202738305 | 0.004542776 | 1.777713049 | IVW |
| 2093 | Knee OA (arcOGEN) | IDP_dMRI_TBSS_ISOVF_Superior_longitudinal_fasciculus_L | 8 | 0.374959486 | 0.18062504 | 0.037903219 | 1.454932468 | IVW |
| 2114 | Knee OA (arcOGEN) | IDP_dMRI_ProbtrackX_ISOVF_ilf_l | 10 | 0.30959031 | 0.141694168 | 0.02889477 | 1.362866647 | IVW |
| 2122 | Knee OA (arcOGEN) | IDP_dMRI_ProbtrackX_ISOVF_slf_r | 18 | 0.288203364 | 0.114741868 | 0.012013275 | 1.33402857 | IVW |
| 2124 | Knee OA (arcOGEN) | IDP_dMRI_ProbtrackX_ISOVF_str_r | 7 | 0.499842719 | 0.229576992 | 0.02946315 | 1.648461979 | IVW |

**BIDPs: Brain imaging-derived phenotypes; SE: standard error; N(SNPs): Number of SNPs; IVW, inverse variance weighted.**

For the IDPs of structural connectivity derived from dMRI data, we found significant causal relationships between parameters on FA (fractional anisotropy) of axonal connections such as the corticospinal tract (CST), the pontine crossing tract (PCT), the superior thalamic radiation, the superior longitudinal fasciculus, the fronto-occipital fasciculus, as well as of local microstructures such as the anterior limb of internal capsule and KOA risk (**Table 1**). Notably, our results supported causal associations between several decreased DTI parameter indicators (including Mean MD, Mean L1/L2/L3, and Mean ISOVF) of CST/PCT and KOA. The detailed MR estimates of the different methods were presented in **Supplementary Table 4**. No directional pleiotropy was found in the MR-Egger regression **(Supplementary Table 5).** The detailed forest plots, scatter plots, funnel plots, and leave-one-out analysis plots for each analysis were presented in **Supplementary Figure 1–50**.

### 3.3 Causal effect of KOA on BIDPs

We further explored the reverse causal effect of KOA on BIDPs using the KOA-related SNPs as IVs, with an *F*-statistic > 10 (range:22.51-28.12, with an average value of 24.91). We found that increased risk of KOA could reduce the thickness of right caudomedial frontal lobe and the area of left parahippocampal gyrus under both Desikan and DKTatlas parcellation (**Table 2**). However, KOA was positively correlated with the thickness and intensity-contrast of the left rostral anterior cingulate, as well as the intensity-contrast of caudal anterior cingulate (left: *β* = 0.071, *p* = 0.017; right: *β* = 0.075, *p* = 0.011), the intensity-contrast of insula cortex in right hemisphere (*β* = 0.072, *p* = 0.016), and the grey matter volume of in both left (*β* = 0.1, *p* = 0.0001) and right (*β* = 0.079, *p* = 0.006) pallidum. In addition, KOA could contribute to decreased Mean ISOVF in medial thalamic (left: *β* = -0.063, *p* = 0.036; right: *β* = -0.064, *p* = 0.033) (**Table 2**).

**Table 2.** The results of Reverse MR (BIDPs were used as outcomes).

| Exposre | Outcome<br>BIDPs<br>ID | IDP short name | KOA vs Control |  |  |  |  |  |
| --- | --- | --- | --- | --- | --- | --- | --- | --- |
| | | | N (SNPs) | $\beta$ | SE | P value | OR | Method |
| Knee OA (arcOGEN) | 18 | IDP_T1_FIRST_right_pallidum_volume | 4 | 0.0807 | 0.0355 | 0.02283319 | 1.084071738 | IVW |
| Knee OA (arcOGEN) | 48 | IDP_T1_FAST_ROIs_L_mid_temp_gyrus_post | 4 | -0.0681 | 0.0291 | 0.01914813 | 0.934167893 | IVW |
| Knee OA (arcOGEN) | 54 | IDP_T1_FAST_ROIs_L_inf_temp_gyrus_post | 4 | -0.0644 | 0.0291 | 0.02670215 | 0.937616755 | IVW |
| Knee OA (arcOGEN) | 70 | IDP_T1_FAST_ROIs_L_latocc_cortex_inf | 4 | -0.0680 | 0.0330 | 0.03922326 | 0.934306506 | IVW |
| Knee OA (arcOGEN) | 94 | IDP_T1_FAST_ROIs_L_parahipp_gyrus_post | 4 | -0.0573 | 0.0291 | 0.04864192 | 0.944294832 | IVW |
| Knee OA (arcOGEN) | 100 | IDP_T1_FAST_ROIs_L_temp_fusif_cortex_post | 4 | -0.0651 | 0.0291 | 0.02518339 | 0.936997495 | IVW |
| Knee OA (arcOGEN) | 120 | IDP_T1_FAST_ROIs_L_occ_pole | 4 | 0.0573 | 0.0291 | 0.0485507 | 1.059016069 | IVW |
| Knee OA (arcOGEN) | 128 | IDP_T1_FAST_ROIs_L_pallidum | 4 | 0.1003 | 0.0301 | 0.000843084 | 1.105519765 | IVW |
| Knee OA (arcOGEN) | 129 | IDP_T1_FAST_ROIs_R_pallidum | 4 | 0.0795 | 0.0291 | 0.006267096 | 1.082702077 | IVW |
| Knee OA (arcOGEN) | 185 | aseg_global_volume_CC-Mid-Anterior | 4 | -0.0645 | 0.0296 | 0.02933929 | 0.937491629 | IVW |
| Knee OA (arcOGEN) | 387 | aparc-Desikan_rh_volume_lateralorbitofrontal | 4 | 0.0635 | 0.0315 | 0.04388895 | 1.065567989 | IVW |
| Knee OA (arcOGEN) | 555 | aparc-a2009s_lh_volume_S-intrapariet+P-trans | 4 | 0.0585 | 0.0296 | 0.0483287 | 1.06023644 | IVW |
| Knee OA (arcOGEN) | 625 | aparc-a2009s_rh_volume_S-front-inf | 4 | -0.0700 | 0.0296 | 0.01813876 | 0.932398762 | IVW |
| Knee OA (arcOGEN) | 663 | aparc-Desikan_lh_area_parahippocampal | 4 | -0.0852 | 0.0296 | 0.004024795 | 0.918324844 | IVW |
| Knee OA (arcOGEN) | 731 | aparc-pial_lh_area_parahippocampal | 4 | -0.0892 | 0.0296 | 0.00260549 | 0.91467058 | IVW |
| Knee OA (arcOGEN) | 794 | BA-exvivo_lh_area_perirhinal | 4 | -0.0586 | 0.0296 | 0.04777777 | 0.94304927 | IVW |
| Knee OA (arcOGEN) | 823 | aparc-DKTatlas_lh_area_parahippocampal | 4 | -0.0856 | 0.0296 | 0.003860497 | 0.917967942 | IVW |
| Knee OA (arcOGEN) | 884 | aparc-a2009s_lh_area_G-front-inf-Orbital | 4 | 0.0631 | 0.0296 | 0.03326742 | 1.06509774 | IVW |
| Knee OA (arcOGEN) | 894 | aparc-a2009s_lh_area_G-oc-temp-med-Parahip | 4 | -0.0621 | 0.0296 | 0.03618754 | 0.93982923 | IVW |
| Knee OA (arcOGEN) | 897 | aparc-a2009s_lh_area_G-pariet-inf-Supramar | 4 | -0.0677 | 0.0296 | 0.022264 | 0.934526458 | IVW |
| Knee OA (arcOGEN) | 917 | aparc-a2009s_lh_area_S-cingul-Marginalis | 4 | 0.0673 | 0.0296 | 0.0231901 | 1.069567969 | IVW |
| Knee OA (arcOGEN) | 1045 | aparc-Desikan_lh_thickness_rostralanteriorcingulate | 4 | 0.0597 | 0.0296 | 0.04393311 | 1.061501961 | IVW |
| Knee OA (arcOGEN) | 1057 | aparc-Desikan_rh_thickness_caudalmiddlefrontal | 4 | -0.0702 | 0.0296 | 0.01783849 | 0.932228577 | IVW |
| Knee OA (arcOGEN) | 1148 | aparc-DKTatlas_rh_thickness_caudalmiddlefrontal | 4 | -0.0736 | 0.0296 | 0.01299452 | 0.929059077 | IVW |
| Knee OA (arcOGEN) | 1306 | aparc-a2009s_rh_thickness_S-interm-prim-Jensen | 4 | -0.0629 | 0.0296 | 0.03377106 | 0.939046801 | IVW |
| Knee OA (arcOGEN) | 1369 | wg_lh_intensity-contrast_caudalanteriorcingulate | 4 | 0.0709 | 0.0296 | 0.0166977 | 1.073473852 | IVW |
| Knee OA (arcOGEN) | 1392 | wg_lh_intensity-contrast_rostralanteriorcingulate | 4 | 0.1038 | 0.0296 | 0.000459863 | 1.109349492 | IVW |
| Knee OA (arcOGEN) | 1404 | wg_rh_intensity-contrast_caudalanteriorcingulate | 4 | 0.0753 | 0.0296 | 0.01108 | 1.078154766 | IVW |
| Knee OA (arcOGEN) | 1436 | wg_rh_intensity-contrast_insula | 4 | 0.0715 | 0.0296 | 0.01574411 | 1.074156369 | IVW |
| Knee OA (arcOGEN) | 1504 | IDP_dMRI_ProbtrackX_FA_cgc_l | 4 | -0.0704 | 0.0299 | 0.0186896 | 0.932053046 | IVW |
| Knee OA (arcOGEN) | 1508 | IDP_dMRI_ProbtrackX_FA_cst_l | 4 | -0.0646 | 0.0299 | 0.0308279 | 0.9374333 | IVW |
| Knee OA (arcOGEN) | 1538 | IDP_dMRI_TBSS_MO_Inferior_cerebellar_peduncle_L | 4 | 0.0690 | 0.0299 | 0.0210732 | 1.071457049 | IVW |
| Knee OA (arcOGEN) | 1551 | IDP_dMRI_TBSS_MO_Superior_corona_radiata_R | 4 | -0.0756 | 0.0299 | 0.01155813 | 0.927222306 | IVW |
| Knee OA (arcOGEN) | 1552 | IDP_dMRI_TBSS_MO_Superior_corona_radiata_L | 4 | -0.0718 | 0.0299 | 0.01642042 | 0.930722233 | IVW |
| Knee OA (arcOGEN) | 1723 | IDP_dMRI_TBSS_L2_Superior_corona_radiata_L | 4 | 0.0705 | 0.0299 | 0.01838723 | 1.073094253 | IVW |
| Knee OA (arcOGEN) | 1879 | IDP_dMRI_ProbtrackX_L3_cgc_l | 4 | 0.0794 | 0.0375 | 0.03446909 | 1.082627189 | IVW |
| Knee OA (arcOGEN) | 2001 | IDP_dMRI_TBSS_OD_Superior_corona_radiata_R | 4 | 0.0653 | 0.0299 | 0.0289965 | 1.067518184 | IVW |
| Knee OA (arcOGEN) | 2002 | IDP_dMRI_TBSS_OD_Superior_corona_radiata_L | 4 | 0.0715 | 0.0299 | 0.01689801 | 1.07409916 | IVW |
| Knee OA (arcOGEN) | 2060 | IDP_dMRI_TBSS_ISOVF_Medial_lemniscus_R | 4 | -0.0639 | 0.0299 | 0.03268759 | 0.938088298 | IVW |
| Knee OA (arcOGEN) | 2061 | IDP_dMRI_TBSS_ISOVF_Medial_lemniscus_L | 4 | -0.0626 | 0.0299 | 0.0364241 | 0.939316581 | IVW |
| Knee OA (arcOGEN) | 2125 | IDP_dMRI_ProbtrackX_ISOVF_unc_l | 4 | 0.0639 | 0.0299 | 0.0327 | 1.0660 | IVW |

**BIDPs: Brain imaging-derived phenotypes; SE: standard error; N(SNPs): Number of SNPs; IVW, inverse variance weighted.**

## 4. Discussion

To the best of our knowledge, this is the first two-sample MR study to investigate causal relationships between KOA and BIDPs. Our study supported bidirectional causal associations between KOA and BIDPs, which may provide new insights into the interaction of KOA with structural alterations in the nervous system.

Although OA and neurological disorders belong to two distinct medical categories, the close association between them has been well documented[5, 6]. A recent MR study further assessed causal relationships between osteoarthritis and central nerve system dysfunction in the elderly, which suggested that there was a positive causal effect of OA on PD, but not on AD nor ischemic stroke[20]. Neuroimaging techniques have been utilized in monitoring the progression and prognosis of neurological diseases. Owing to the increasing availability of MRI-based GWAS, structural imaging studies have revealed alterations in BIDPs in neurological disorders. For example, the causal AD-associations for the left hippocampus and right inferior temporal cortex volumes have been identified by utilizing brain MRI database from the Alzheimer’s Disease Neuroimaging Initiative and the UK Biobank[21]. However, the causal relationship between BIDP alterations and KOA is still unclear.

Our findings supported that increased area of the paracentral lobule (PCL) could reduce the risk of KOA. The PCL is on the medial surface of the cerebral hemisphere and is the continuation of the precentral and postcentral gyri. In the human brain, motor control and somatosensory representations of different parts of the body are ordered respectively along the entire range of the precentral and postcentral gyrus (generally categorized into five sections: toes/legs, arms/hands, blinking, mouth, and tongue)[22, 23]. Interestingly, the PCL controls motor and sensory innervations of the contralateral lower extremity, including the knee. Pain is an important somatosensation and a major symptom of OA, especially, in KOA where pain is recognized as typically changing from intermittent weight-bearing pain to chronic pain[24]. Several studies have reported the association between chronic pain and reduced gray matter in areas associated with pain processing and modulation, such as the dorsolateral prefrontal cortex, thalamus, dorsolateral pons, and somatosensory cortex[25–27]. Another study also reported a significant reduction in gray matter volume in the left precentral gyrus in patients with KOA relative to healthy controls[28]. Here we identified that alterations in the area of PCL as a cause, rather than a consequence, of KOA. Moreover, we found that increased BA4a area in the left hemisphere was also a protective factor for KOA. BA4 belongs to the anterior two-thirds of the PCL (medial surface of the precentral gyrus), where the ungranulated cortex reflects the primary motor cortex in the PCL for the contralateral leg and foot muscles, reinforcing its possible role in the pathogenesis of OA[29].

The nucleus accumbens (NAc) is a subcortical brain structure located in the ventral striatum. It is often known as the pleasure center and is closely associated with behaviors such as reward, motivation, and learning[30]. Its imbalance is commonly associated with many mental and neurological disorders such as depression, obsessive-compulsive disorder, anxiety, and substance abuse[31]. In recent years, however, the NAc has been shown to play a role in acute and chronic pain modulation[32, 33], and even to be effective in pain relief through deep brain stimulation of the human NAc[34]. Although the current study found a causal link between increased volume but decreased intensity-contrast of NAc and KOA risk, further studies are still needed to determine whether the altered neuroimaging in this region is associated with changes in the size, number, and function of neurons or glial cells.

DTI quantifies the Brownian motion of water molecules in tissues and organs, which is sensitive in detecting the integrity and direction of fiber bundles[35]. Considered to be the most useful parameter of DTI, FA reflects the integrity of myelin sheaths as well as the number, size, and density of fibers. Pathological process impairing the microstructure of the CNS (such as inflammation, demyelination, or axonal degeneration) will alter the anisotropy of water diffusion and thus the FA. CST transmit output signals of superficial neurons and ultimately projects to the spinal cord anterior horn cells to regulate basic motor functions[36]. As the most common motor neuron disease, amyotrophic lateral sclerosis (ALS), there have been extensive studies showing that the FA of the CST was reduced and correlates with functional scores as well as progression in ALS patients[37–39]. In the current study, we confirmed a causal link between decreased DTI parameters on FA in CST and KOA risk. Although the underlying mechanisms are unknown, these indicators may provide new perspectives for the screening of KOA susceptibility in the future.

On the other hand, the current findings suggest that KOA in turn may also lead to an increment of thickness or intensity-contrast in the cingulate, insula, and pallidum. Reduced gray matter volume or density often corresponds to neuronal degeneration or death, as manifested in patients with cerebral hypoxia or infarction[40, 41], and conversely we believe it may be related to neural sensitization. Peripheral and central sensitization has been demonstrated in patients with OA, which leads to local nociceptive sensitization as manifested by decreased pain thresholds[42]. Our results are consistent with a previous pain fMRI meta-analysis study indicating that patients with chronic pain, or experimental pain caused by nociceptive hypersensitivity, showed stronger activation in these brain regions[43]. Moreover, the cingulate and insula are important component of the limbic system, which has a key role in coordinating injurious, cognitive-emotional pain processing[44, 45]. It has also been established that depression or low mood triggered by chronic pain contribute to central pain exacerbation and severity[46, 47]. Our findings may provide new explanations for nociceptive hypersensitivity of chronic pain in KOA patients.

This study is not without limitations. First, the study used GWAS summary data, which may not allow for stratified analysis of relevant risk factors such as gender, age, obesity, and OA severity. Second, the data of exposure and outcome were obtained only from European populations, which limits the cross-ethnic extrapolation of our findings. Third, as the function of many brain regions is not yet fully understood, which constrained the interpretation of our MR results. In conclusion, this study demonstrated a bidirectional causal relationship between BIDPs and KOA risk. Further studies are needed to confirm and further explore these findings.

## Data Availability

https://open.win.ox.ac.uk/ukbiobank/big40

## Author contribution Statement

S.-S.Z. and H.-T.L.designed this study. Z.R. and S.-H.L.wrote the manuscript. S.-S.Z., Z.R. and S.-H.L analyzed the data and prepared all the figures. C.P., T.-T.X., and L.M. provided technical support. H.- T.L., and S.-S.Z. provided financial support.

## Ethics approval

All analyses were based on publicly shared databases, and no additional ethical approvals were required.

## Conflict of Interest Statement

No conflict of interest declared.

## Funding

This work was financially supported by the National Natural Science Foundation of China (Nos. 81902222, and Nos.82060395).

## Data Availability Statement

All the data are available if qualified authors apply for them.

## Notes

### Competing Interest Statement

NO authors have competing interests

### Funding Statement

Yes

